# A Generalized Multinomial Probabilistic Model for SARS-CoV-2 Infection Prediction and Public Health Intervention Assessment in an Indoor Environment

**DOI:** 10.1101/2022.12.02.22282697

**Authors:** Victor OK Li, Jacqueline CK Lam, Yuxuan Sun, Yang Han, Kelvin Chan, Shan-shan Wang, Jon Crowcroft, Jocelyn Downey, Qi Zhang

## Abstract

SARS-CoV-2 Omicron has become the predominant variant globally. Current infection models are limited by the need for large datasets or calibration to specific contexts, making them difficult to cater for different settings. To ensure public health decision-makers can easily consider different public health interventions (PHIs) over a wide range of scenarios, we propose a generalized multinomial probabilistic model of airborne infection to systematically capture group characteristics, epidemiology, viral loads, social activities, environmental conditions, and PHIs, with assumptions made on social distancing and contact duration, and estimate infectivity over short time-span group gatherings. This study is related to our 2021 work published in Nature Scientific Reports that modelled airborne SARS-CoV-2 infection (Han, Lam, Li, et al., 2021).^1^ It is differentiated from former works on probabilistic infection modelling in terms of the following: (1) predicting new cases arising from more than one infectious in a gathering, (2) incorporating additional key infection factors, and (3) evaluating the effectiveness of multiple PHIs on SARS-CoV-2 infection simultaneously. Although our results reveal that limiting group size has an impact on infection, improving ventilation has a much greater positive health impact. Our model is versatile and can flexibly accommodate other scenarios by allowing new factors to be added, to support public health decision-making.

## 1. Introduction

The SARS-CoV-2 Omicron variant, first detected in November 2021 in Botswana, has now become the globally dominant variant.^2^ Omicron carries more than 30 mutations in the spike glycoprotein and evades over 85% of tested neutralizing antibodies,^3^ resulting in a higher risk of reinfection due to its greater ability to evade immunity from prior infection and reduced vaccine effectiveness.^4^

SARS-CoV-2 variants are primarily transmitted through close interpersonal contact.^5^ Non-clinical public health interventions (PHI), such as social distancing and lockdowns, have been introduced in many countries to reduce transmission,^6^ but have met with varying degrees of success in implementation and enforcement^7^ and carry economic and social costs. To inform public health decisions on the type and timing of PHIs, we have sought to create a simple, comprehensive and versatile SARS-CoV-2 infection model that can estimate infections over a wide range of scenarios and be easily updated to reflect changing key empirical characteristics of SARS-CoV-2, its modes and rate of transmission, and public health measures.

### Research Gaps and Aims of the Study

Current models of SARS-CoV-2 transmission primarily rely on viral reproduction rate and Susceptible-Infected-Recovered (SIR) / Susceptible-Exposed-Infectious-Recovered (SEIR) transmission frameworks/models.^8^ These models compute the instantaneous reproductive numbers, parameterize the contact matrix, and then fit the models to epidemic curves. This approach is able to estimate infections across a large population group but requires infection case data over a longer time span (i.e. several days). In the real-world, data limitations may arise, especially over short time spans – for example, data may be delayed, inaccurate, incomplete or even not obtainable at all – and result in a potentially severe loss of accuracy. ^9^

In contrast, other SARS-COV-2 infection models that statistically estimate viral infection risks are often context-based and lack generalizability. For example, Moritz *et al*. (2021) developed a transmission model based on data from an experimental indoor mass gathering and computational fluid dynamics model (CFD) simulations^10^ and Singanayagam *et al*. (2022) estimated the risk of community-based transmission in the United Kingdom.^11^ However, their reference to specific contexts/scenarios restricts their transferability to other settings and CFD models in particular are highly geometrically-specific and determining distribution values across different room settings can be very time-consuming. Additionally, both studies were relatively limited in providing a comprehensive and generalizable account of the risk factors affecting SARS-CoV-2 transmission/infection in different contexts.

To date, few probabilistic models have been developed to uncover the transmission patterns of SARS-CoV-2, especially for small group activities. One notable example is Tupper *et al*. (2020),^12^ which proposed a simple mathematical model of transmission rate based on social contact (contact rate, duration, and mixing level) and calibrated this for different outbreaks. However, the model was limited to new infections arising from a single infectious individual only and did not incorporate a broader range of biological, behavioral, and environmental risk factors affecting infection in different scenarios, such as variant infectivity, vaccination or ventilation conditions.

In this study, we propose a multinomial probabilistic model to predict infections arising from short time span group activities. The model can comprehensively capture key factors affecting SAR-CoV-2 infection; be adapted to model different scenarios; and incorporate additional factors affecting transmissibility as research and circumstances develop.

### Factors Affecting Contacts

Close and prolonged indoor contact can increase viral transmission of SARS-CoV-2.^13^ The longer an infectious person remains in close contact with others, the more likely an exposed (or uninfected) person is to become infected. Similarly, more infectious people or more people in a room increases the number of close contacts between infectious persons and exposed persons and the number of likely infections.

### Factors Affecting Transmissibility

SARS-CoV-2 infection requires a relatively low initial infectious dose compared to other respiratory viruses^14^. Multiple factors affect transmissibility, including: the infectivity of the viral variant, the contagiousness of infected individuals, the susceptibility of exposed individuals, the nature of contact, the contact environment, and any infection reduction measures undertaken.^11,14,15^

At the biological level, despite the virology and physiology of SARS-CoV-2 are yet to be fully understood, several papers have noted differences in infectivity between SARS-CoV-2 variants.^16-18^ Thus, Blanquart *et al*. (2021) observed a difference in viral loads between vaccinated and unvaccinated individuals for Delta, though this was statistically significant only for asymptomatic infections.^19^ The effectiveness of vaccinations against emerging variants is the subject of ongoing investigation ^11,18,20-23^ but studies, such as Levine-Tiefenbrum *et al*. (2021), suggest lower viral loads during breakthrough infections for at least some types of vaccine.^20^

At the behavioral level, physical proximity and duration of exposure influence the probability of infection,^10,24,25,10,24^ as does the activity undertaken – for instance, singing produces a significantly higher viral load than normal breathing.^15,26,27^ Wearing face masks or using shielding can reduce the risk of infection,^10,28-32^ with effectiveness depending inter alia on mask material and compliance.^30,33^

At the environmental level, ventilation plays a critical role in air-borne SARS-CoV-2 transmission^10,15,34-36^ by directing, dispersing and diluting expelled infectious droplets/aerosols.^37^ Outdoor settings with better natural air flows than indoor spaces can further reduce airborne viral transmission.^38^

There are likely further risk factors. In particular, demographic influences are believed to be significant in determining infection outcomes,^39,40^ but since the relationship between demographic factors and of SARS-CoV-2 infection risk has not been fully quantified at the time of writing this paper, the proposed model does not take demographic factors into account.

### Research Questions

Firstly, given M infectious persons and a total of N persons (N-M exposed persons) in a room, we consider the number of exposed persons who are expected to become infected in a time period T. Secondly, we examine how partitioning a large group into smaller groups affects the average number of exposed persons becoming infected. Thirdly, we investigate how the probability of becoming infected is impacted by changes in each of the following conditions: the activity of the infectious persons and of the exposed persons (including breathing only, speaking, etc.), masking, vaccination, ventilation, viral load, and variant infectivity.

### Novelties and Significance of Study

The proposed model has a simple and versatile multinomial probabilistic structure that can systematically capture known infection risk factors, including: group characteristics (group size); biological factors (infectivity of the variant, vaccinated/unvaccinated); human behaviours (duration of exposure, activities undertaken, masked/unmasked); and environmental conditions (ventilation). The model is readily extended to include additional epidemiological risk factors and its bottom-up structure allows a wide range of scenarios to be considered. Compared to earlier probabilistic models, the proposed model is more comprehensive, capturing cases with one or multiple infectious persons and incorporating more key risk factors affecting transmission and infection.

Our proposed model determines expected infection cases under different circumstances, offering, for example, quick and useful public health guidance on how to reduce infections during large group gatherings. This is particularly important at the present time as cities increasingly seek to end social distancing policies and resume normal daily social and economic activities as much as possible, while minimizing the risks of large-scale infection and the resultant adverse health consequences.

## 2. Results

### 2.1 Basic model: M infectious persons in one single group gathering of N persons

Suppose there are N persons in a room with M infectious persons, i.e., there are N-M exposed (healthy) persons, with each infectious person making L contacts at random in a time period T, and infecting each exposed (healthy) person, with probability p for each contact. We are interested in I, the number of exposed persons newly infected. The formulae derived in Eq(1) and Eq(2) in the Methods section give the distribution of I and E(I), the expected average of I, respectively.

Figure 1a shows the average number of infections E(I) for group size N ranging from 1 to 100, for time duration (in hours) T = 1, 2, 3, and 4, and for p = 1. Here we have assumed that M = 1, i.e., there is only one infectious person, and that this infectious person will make one contact every 15 minutes. As expected, the average number of infected persons increases with the number of persons in the room and with T, until E(I) reaches a saturation point. In Figure 1a, for each T, the 10% saturation point is indicated with a diamond and the 5% saturation point is indicated with a circle. A 10% saturation point means the lowest N such that when N increases by 1, E(I) increases by less than 10%. Similarly, a 5% saturation point means the lowest N such that when N increases by 1, E(I) increases by less than 5%. When T = 1 (L = 4), 10% and 5% saturation occur at N = 9 and 11, respectively; when T = 4 (L = 16), 10% and 5% saturation points occur at N = 32 and 44, respectively. Table 1a shows the average number of infected persons when the size of the group increases from 50 to 500 for time durations from one to four hours.

**Figure 1a.**
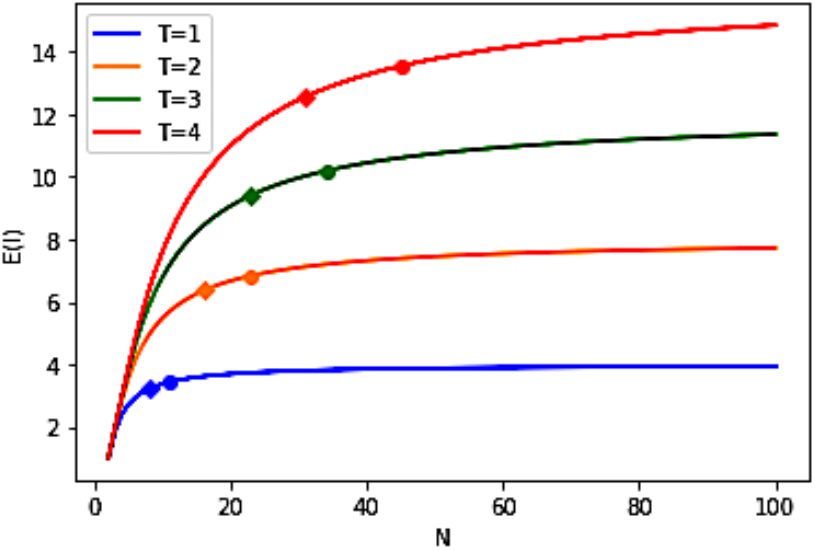
E(I) vs N, for T = 1, 2, 3, and 4, M=1, and p =1 (Diamond indicates 10% saturation and circle indicates 5% saturation for a given curve).

**Figure 1b.**
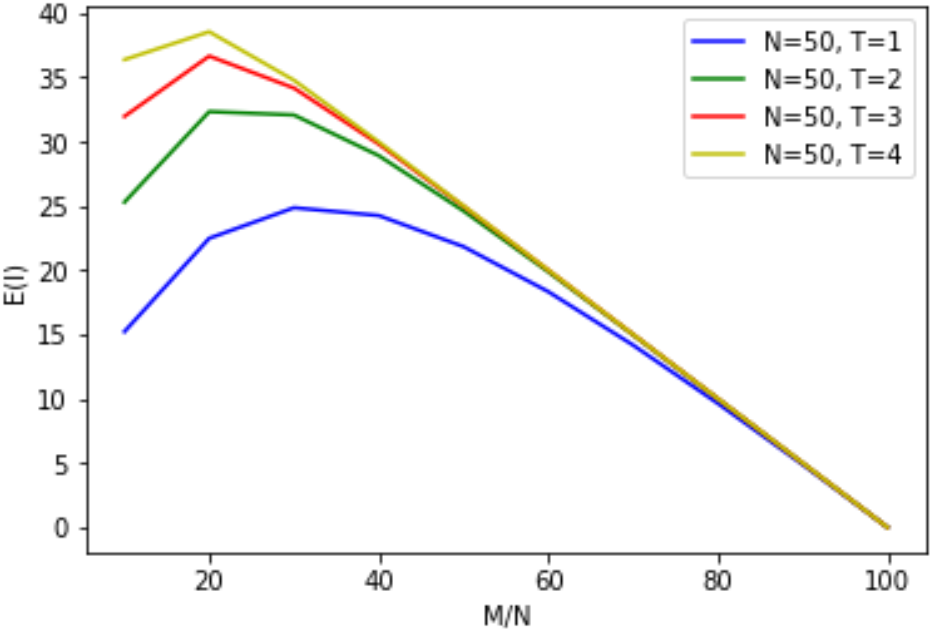
E(I) vs M/N, for T = 1, 2, 3, and 4, N=50, and p =1.

**Figure 1c.**
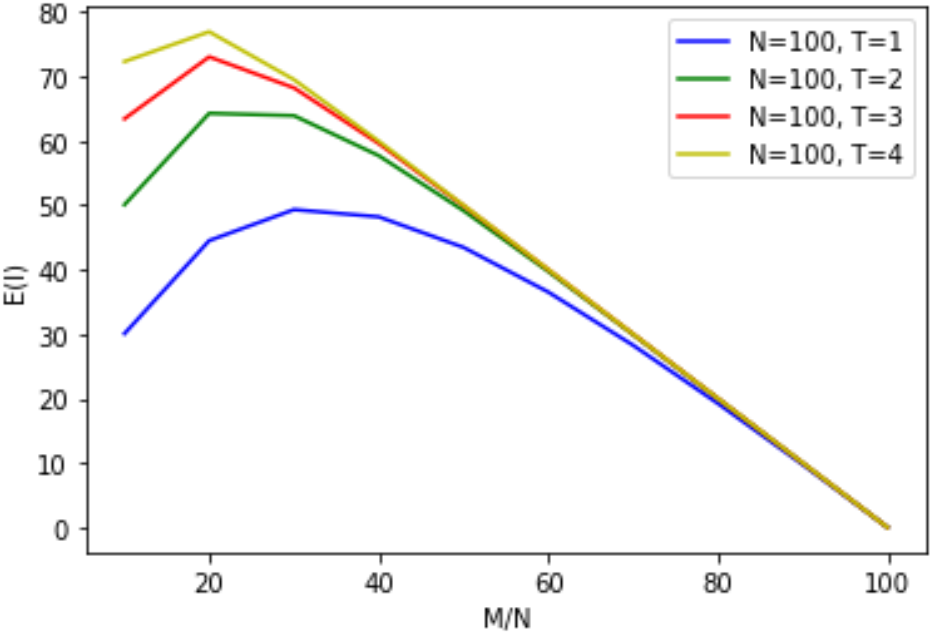
E(I) vs M/N, for T = 1, 2, 3, and 4, N=100, and p =1.

**Figure 1d.**
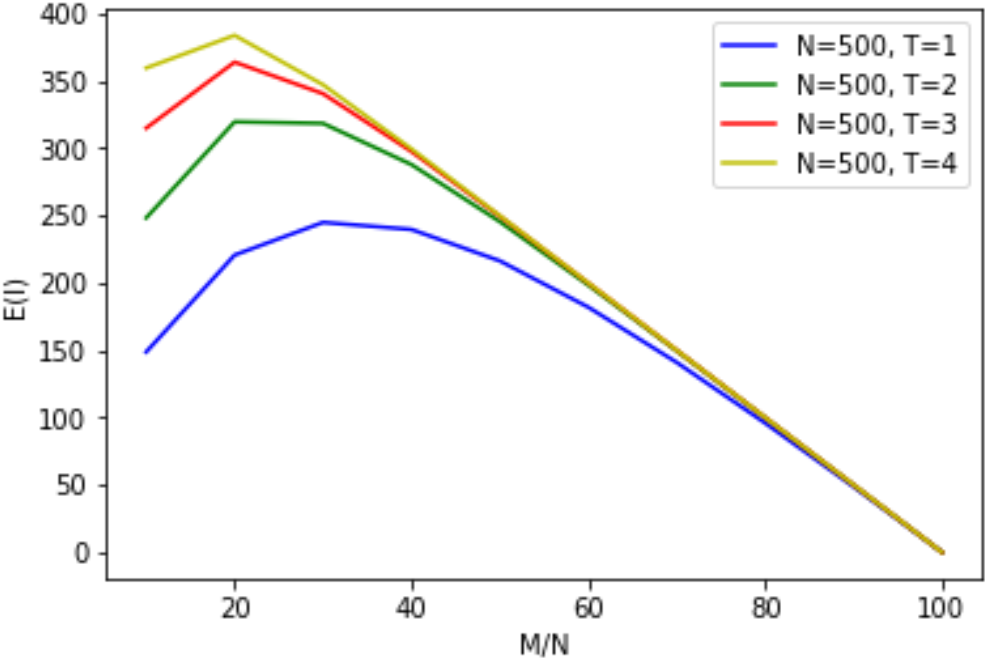
E(I) vs M/N, for T = 1, 2, 3, and 4, N=500, and p =1.

**Table 1a.** E(I) vs N, when N = 50, 100, and 500, for T = 1, 2, 3, 4, M=1, and p =1.

| No. of People in a Room ($N$) | $E(I)$ | | | |
| --- | --- | --- | --- | --- |
| | $T = 1$ | $T = 2$ | $T = 3$ | $T = 4$ |
| **50** | 3.88 | 7.45 | 10.74 | 13.77 |
| **100** | 3.94 | 7.72 | 11.36 | 14.84 |
| **500** | 3.99 | 7.94 | 11.87 | 15.76 |

**Table 1b.** E(I) vs M/N, when N = 50, 100, and 500, for M/N=20% and 30%, T = 1, 2, 3, 4, M=1, and p =1.

| $M/N$ vs $N$ | $T=1$ | $T=2$ | $T=3$ | $T=4$ |
| --- | --- | --- | --- | --- |
| **$N=50$; $M/N=20\%$** | 22.47 | 32.31 | 36.63 | 38.52 |
| **$N=100$; $M/N=20\%$** | 44.49 | 64.24 | 73.00 | 76.89 |
| **$N=500$; $M/N=20\%$** | 220.70 | 319.63 | 363.97 | 383.85 |
| **$N=50$; $M/N=30\%$** | 24.84 | 32.05 | 34.14 | 34.75 |
| **$N=100$; $M/N=30\%$** | 49.30 | 63.88 | 68.19 | 69.46 |
| **$N=500$; $M/N=30\%$** | 244.96 | 318.48 | 340.54 | 347.16 |

Table 1b and Figure 1b-1d show that given the same duration T, as M/N increases, the number of new cases, E(I), due to incoming participants to the room who are infectious, also increases, until it reaches a peak. For T = 1, this peak occurs at M/N = 30% for N = 50, 100, and 500. For T = 2, 3, and 4, this peak occurs at M/N = 20% for N = 50, 100, and 500. Subsequent to the peak, E(I) decreases, as the number of exposed becomes smaller as M/N increases. When M/N is 100%, there are no exposed persons to be infected. Given the same group size, N, and an infectious percentage, M/N, a smaller T (e.g., T=1) always produces a lower infection number, E(I), as compared to a larger T (e.g., T=2, 3, 4).

### 2.2 Partition model: M infectious persons in one single indoor meeting with N persons partitioned at random into multiple rooms

We consider three different group gathering sizes, mimicking different real world social settings, such as classrooms, social gatherings, and conferences. The formulae derived in the Methods section allow us to determine the average number of persons who become infected when a single group gathering of size N with M infectious persons is partitioned at random into smaller groups and placed in separate rooms.

Consider a gathering of size N = 50 and the cases where infectious persons number M = 5, 10, and 15 amongst these 50 persons, i.e., M/N = 10%, 20%, and 30% respectively; as all individuals are in one room, we consider this as partition size 1, i.e. PAR = 1. Next, repeat but randomly partition the group of N = 50 into two separate rooms of 25, i.e. PAR = 2, and similarly for five separate rooms of 10, i.e. PAR = 5. Finally, for each case calculate E(I), the expected number of persons who become infected. The results, together with those of larger groups N = 100 and N = 500, are shown in Table 2 (for p = 1) and demonstrate that partitioning has a very limited effect in reducing E(I). The greatest reduction in E(I) is achieved when the percentage of infectious persons M/N is 30% (Figure 2). E(I) as a proportion of N declines slightly as group size N increases but this is highly constrained. Details of the methodology can be found in the Methods section, for the case p = 1.

**Table 2.** The expected number of people becoming infected E(I) for p = 1, T=1, M/N = 10%, 20% …, 100%, N = 50, 100, 500 and PAR = 1, 2, 5.

| | No. of Partitions (PAR) | Partition Size | E(I) for $p = 1$ | | | | | | | | | |
| --- | --- | --- | --- | --- | --- | --- | --- | --- | --- | --- | --- | --- |
|  |  |  | % Infectious (M/N) |  |  |  |  |  |  |  |  |  |
|  |  |  | 10% | 20% | 30% | 40% | 50% | 60% | 70% | 80% | 90% | 100% |
| Small Group Gathering<br>$N = 50$ | 1 | 50 | 15.21 | 22.47 | **24.84** | 24.24 | 21.82 | 18.32 | 14.16 | 9.63 | 4.88 | 0 |
|  | 2 | 25 | 14.78 | 21.92 | **24.32** | 23.79 | 21.46 | 18.05 | 13.98 | 9.55 | 4.87 | 0 |
|  | 5 | 10 | 13.45 | 20.20 | **22.65** | 22.37 | 20.40 | 17.37 | 13.66 | 9.45 | 4.86 | 0 |
| Medium Group Gathering<br>$N = 100$ | 1 | 100 | 30.04 | 44.49 | **49.30** | 48.18 | 43.44 | 36.50 | 28.25 | 19.22 | 9.74 | 0 |
|  | 2 | 50 | 29.63 | 43.96 | **48.79** | 47.75 | 43.09 | 36.24 | 28.05 | 19.10 | 9.71 | 0 |
|  | 5 | 20 | 28.38 | 42.35 | **47.23** | 46.40 | 42.02 | 35.46 | 27.61 | 18.94 | 9.69 | 0 |
| Large Group Gathering<br>$N = 500$ | 1 | 500 | 148.72 | 220.70 | **244.96** | 239.72 | 216.37 | 181.99 | 140.96 | 95.96 | 48.65 | 0 |
|  | 2 | 250 | 148.33 | 220.19 | **244.47** | 239.30 | 216.03 | 181.72 | 140.76 | 95.82 | 48.56 | 0 |
|  | 5 | 100 | 147.14 | 218.66 | **242.99** | 238.02 | 215.00 | 180.93 | 140.17 | 95.42 | 48.43 | 0 |

**Figure 2.**
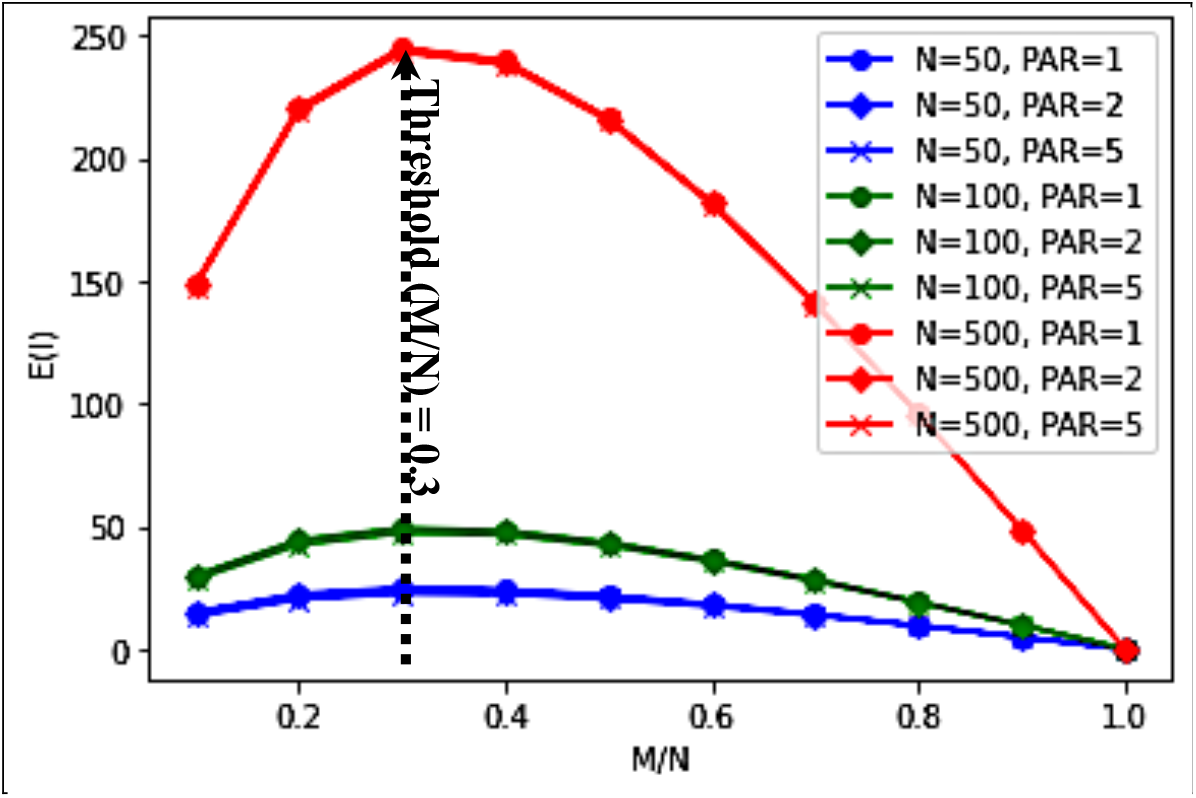
The expected number of people becoming infected for p = 1, T=1, M/N = 10%, 20%, …, 100%, N = 50, 100, 500 and PAR = 1, 2, 5.

### 2.3 Average number of people getting infected in an indoor setting under different values of infection probability p

Our model allows us to explore how E(I) changes for different values of infection probability p, reflecting different social, environmental, and public health measures. Tables 3a-3b and Figures 3a-3b show the expected number of people infected for different group size N, partition size PAR, infection probability p and ratios of infectious persons to total group size M/N, when the duration of the gathering is 1 hour (T = 1). Peak E(I)/N is relatively insensitive to group size and number of partitions. Peak E(I) is [moderately] sensitive to M/N: for N = 100, PAR = 1, peak E(I) occurs at M/N = 50% for p = 0.1 and p = 0.3, M/N = 30% for p = 0.5 and p = 1. The rate of change in E(I) peaks when p = 0.5, for all group sizes and all number of partitions.

**Table 3a.**
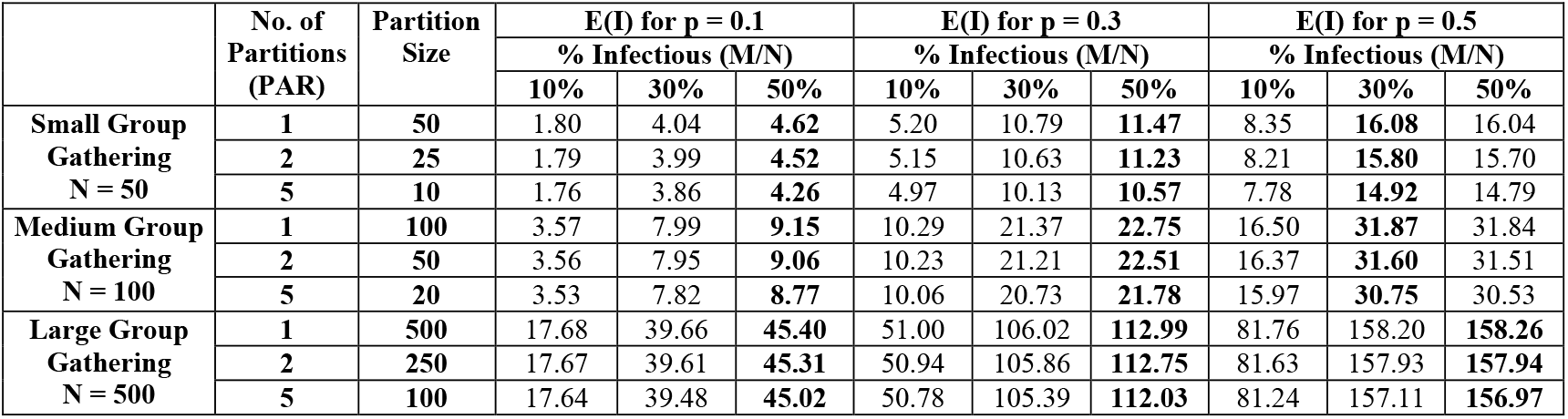
The expected number of people infected E(I) for p = 0.1, 0.3, 0.5; T = 1; M/N = 10%, 30%, 50%; N = 50, 100, 500 and PAR = 1, 2, 5.

**Table 3b.**
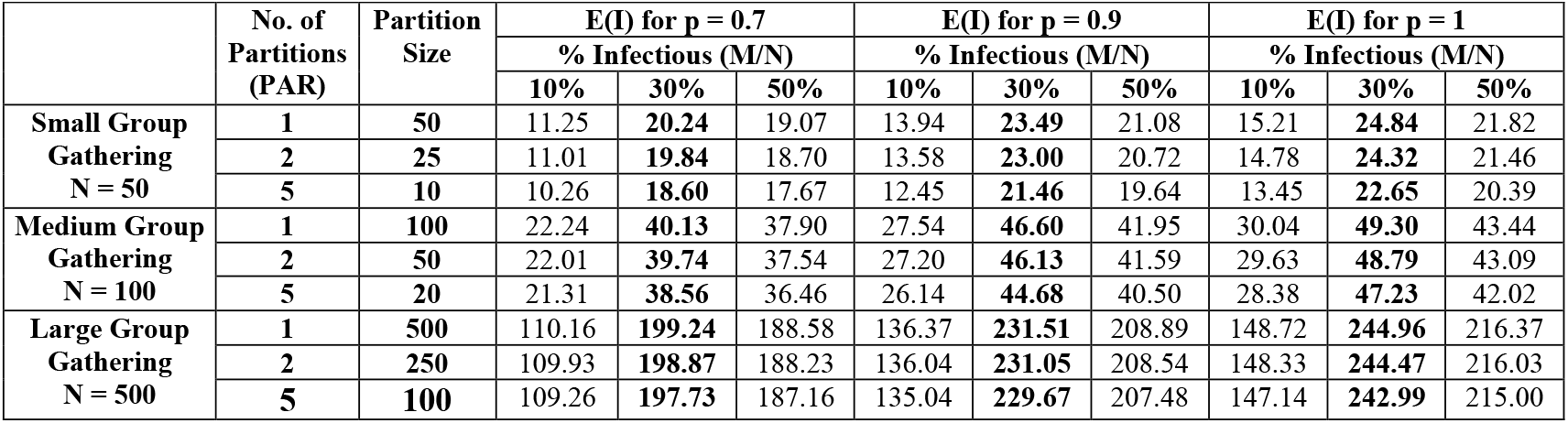
Table 3b. The expected number of people infected E(I) for p = 0.7, 0.9, 1.0, T = 1, M/N = 10%, 30%, 50%, N = 50, 100, 500 and PAR = 1, 2, 5.

| | No. of Partitions (PAR) | Partition Size | E(I) for $p = 0.7$ | | | E(I) for $p = 0.9$ | | | E(I) for $p = 1$ | | |
| --- | --- | --- | --- | --- | --- | --- | --- | --- | --- | --- | --- |
|  |  |  | % Infectious (M/N) |  |  | % Infectious (M/N) |  |  | % Infectious (M/N) |  |  |
|  |  |  | 10% | 30% | 50% | 10% | 30% | 50% | 10% | 30% | 50% |
| Small Group Gathering<br>N = 50 | 1 | 50 | 11.25 | **20.24** | 19.07 | 13.94 | **23.49** | 21.08 | 15.21 | **24.84** | 21.82 |
|  | 2 | 25 | 11.01 | **19.84** | 18.70 | 13.58 | **23.00** | 20.72 | 14.78 | **24.32** | 21.46 |
|  | 5 | 10 | 10.26 | **18.60** | 17.67 | 12.45 | **21.46** | 19.64 | 13.45 | **22.65** | 20.39 |
| Medium Group Gathering<br>N = 100 | 1 | 100 | 22.24 | **40.13** | 37.90 | 27.54 | **46.60** | 41.95 | 30.04 | **49.30** | 43.44 |
|  | 2 | 50 | 22.01 | **39.74** | 37.54 | 27.20 | **46.13** | 41.59 | 29.63 | **48.79** | 43.09 |
|  | 5 | 20 | 21.31 | **38.56** | 36.46 | 26.14 | **44.68** | 40.50 | 28.38 | **47.23** | 42.02 |
| Large Group Gathering<br>N = 500 | 1 | 500 | 110.16 | **199.24** | 188.58 | 136.37 | **231.51** | 208.89 | 148.72 | **244.96** | 216.37 |
|  | 2 | 250 | 109.93 | **198.87** | 188.23 | 136.04 | **231.05** | 208.54 | 148.33 | **244.47** | 216.03 |
|  | 5 | 100 | 109.26 | **197.73** | 187.16 | 135.04 | **229.67** | 207.48 | 147.14 | **242.99** | 215.00 |

**Figure 3a.**
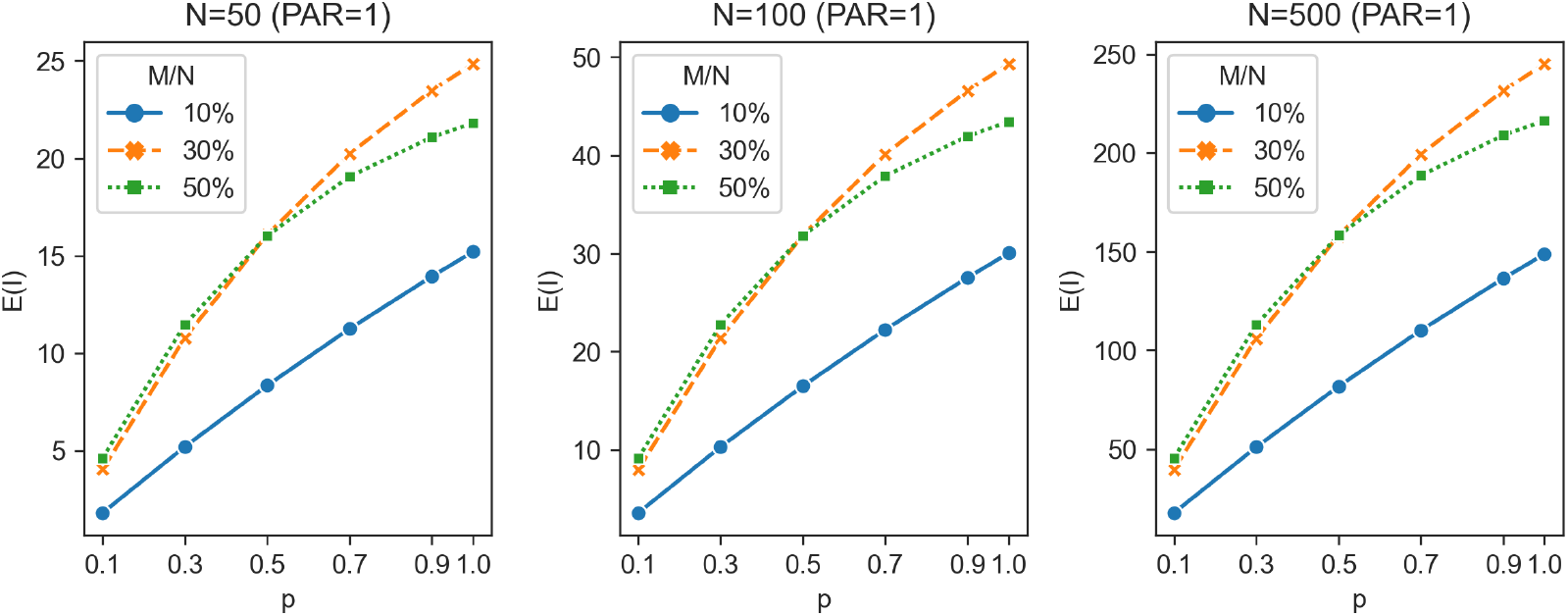
The expected number of people infected E(I) for p = 0.1, 0.3, 0.5, 0.7, 0.9, 1.0 (y-axis), under T = 1; group size N = 50, 100, 500; PAR = 1; M/N = 10%, 30%, 50%

**Figure 3b.**
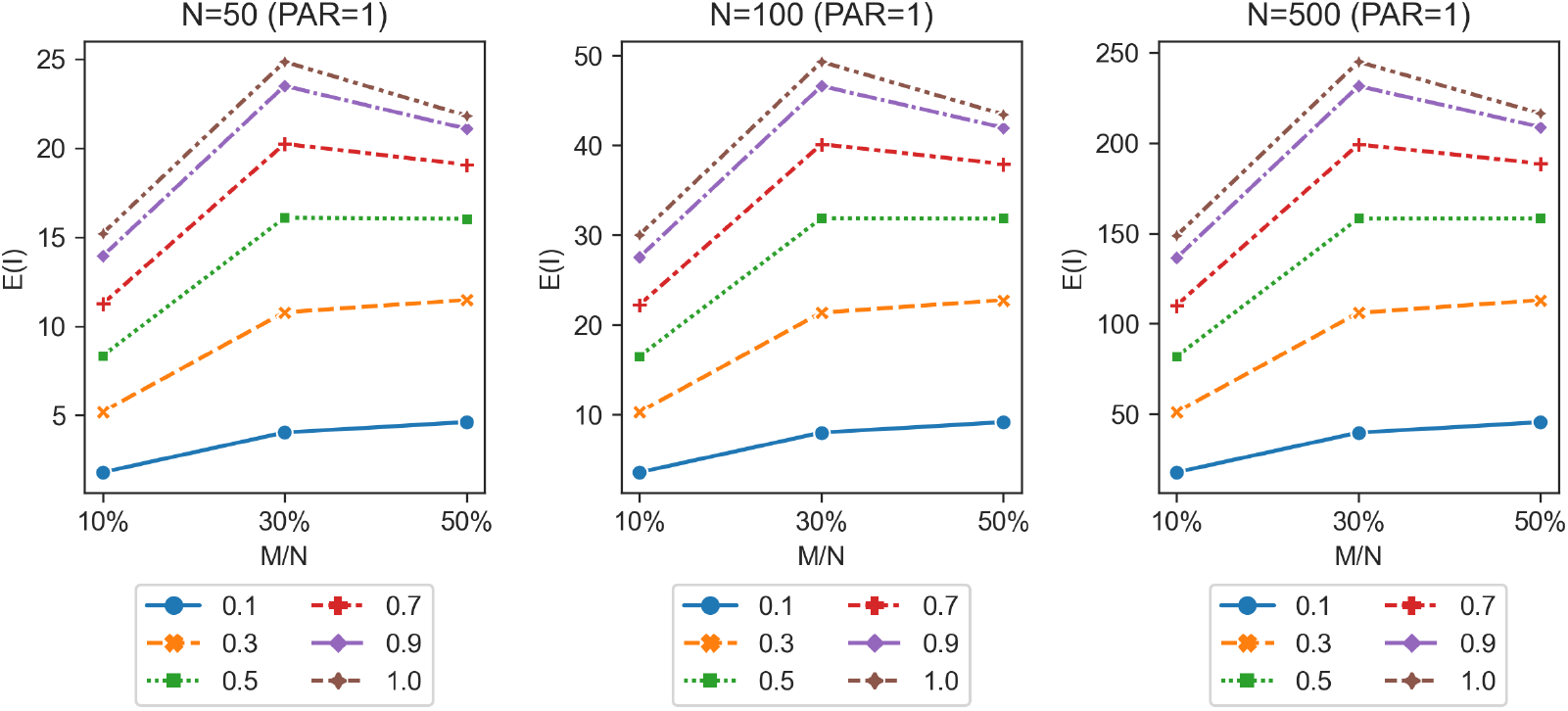
The expected number of people infected E(I) for M/N = 10%, 30%, 50% (y-axis), under T = 1; group size N = 50, 100, 500; PAR = 1; p = 0.1, 0.3, 0.5, 0.7, 0.9

### 2.4 The probability of infection p under different interaction activities and public health interventions

Table 4a shows the probability of infection p under four different indoor “activities”, namely: (1) neither exposed persons nor infectious persons speak (i.e. they only breathe passively), (2) exposed persons do not speak but infectious persons do, (3) exposed persons speak but infectious persons do not speak, and (4) both exposed persons and infectious persons speak. Each scenario is considered under 8 different public health interventions (PHIs), reflecting different parameter values for Masking Factor (MF), Vaccination Factor (VAF), Ventilation Factor (VEF), and Variant Infectivity Factor (CVF). For each of the resultant 32 scenarios, the probability of infection, p, and the relative reduction in p (i.e. the ratio of p under that particular scenario to p under the applicable baseline scenario), p-ratio, are shown. For each activity, the first row shows the baseline scenario without any public health intervention, namely all persons do not wear masks (MF = 1), all persons have not been vaccinated (VAF = 1), the room has minimum mechanical ventilation (VEF = 1), the variant is Omicron (CVF = 1), and one “quanta” of the virus is inhaled (Quantum = 1). Subsequent rows reflect different PHIs and combinations of PHI, with scenarios *h* and *k* reflecting a hypothetical new SARS-CoV-2 variant that is 10x more transmissible than Omicron.

**Table 4a.** The p-ratio and reduction in p for different situations and public health interventions

| **1. Scenario 1: Exposed Persons and Infectious Persons – Both No Speaking** |  |  |  |  |  |  |  |  |  |  |
| --- | --- | --- | --- | --- | --- | --- | --- | --- | --- | --- |
| **Public Health Interventions (PHIs)** | **AFE** | **AFI** | **MF** | **VAF** | **VEF** | **CVF** | **Viral Load (Quanta)** | **p** | **p-ratio** | **Reduction in p due to PHIs** |
| 1a. Baseline | 1.00 | 1.00 | 1.00 | 1.00 | 1.00 | 1.00 | 1.00 | 0.63 | 1.00 |  |
| 1b. Mask | 1.00 | 1.00 | 0.41 | 1.00 | 1.00 | 1.00 | 0.41 | 0.34 | 0.53 | 47.00% |
| 1c. Vac | 1.00 | 1.00 | 1.00 | 0.64 | 1.00 | 1.00 | 0.64 | 0.47 | 0.75 | 25.47% |
| 1d. MaxMV | 1.00 | 1.00 | 1.00 | 1.00 | 0.31 | 1.00 | 0.31 | 0.27 | 0.42 | 57.83% |
| 1e. Mask+Vac+MaxMV | 1.00 | 1.00 | 0.41 | 0.64 | 0.31 | 1.00 | 0.08 | 0.08 | 0.12 | 87.75% |
| 1f. Mask+Vac+MedMV | 1.00 | 1.00 | 0.41 | 0.64 | 0.51 | 1.00 | 0.13 | 0.12 | 0.20 | 80.40% |
| 1g. Mask+Vac+NatV | 1.00 | 1.00 | 0.41 | 0.64 | 0.82 | 1.00 | 0.21 | 0.19 | 0.30 | 69.70% |
| 1h. Mask+Vac+MaxMV+NewVariant | 1.00 | 1.00 | 0.41 | 0.64 | 0.31 | 10.00 | 0.81 | 0.55 | 0.88 | 12.48% |
| 1i. Outdoor | 1.00 | 1.00 | 1.00 | 1.00 | 0.03 | 1.00 | 0.03 | 0.03 | 0.05 | 94.65% |
| 1j. Mask+Vac+Outdoor | 1.00 | 1.00 | 0.41 | 0.64 | 0.03 | 1.00 | 0.01 | 0.01 | 0.01 | 98.59% |
| 1k. Mask+Vac+Outdoor+NewVariant | 1.00 | 1.00 | 0.41 | 0.64 | 0.03 | 10.00 | 0.09 | 0.09 | 0.14 | 86.47% |
| **2. Scenario 2: Exposed Persons No Speaking and Infectious Persons Speaking** |  |  |  |  |  |  |  |  |  |  |
| **Public Health Interventions (PHIs)** | **AFE** | **AFI** | **MF** | **VAF** | **VEF** | **CVF** | **Viral Load (Quanta)** | **p** | **p-ratio** | **Reduction in p due to PHIs** |
| 2a. Baseline | 1.00 | 12.90 | 1.00 | 1.00 | 1.00 | 1.00 | 12.90 | 1.00 | 1.00 |  |
| 2b. Mask | 1.00 | 12.90 | 0.41 | 1.00 | 1.00 | 1.00 | 5.26 | 0.99 | 0.99 | 0.52% |
| 2c. Vac | 1.00 | 12.90 | 1.00 | 0.64 | 1.00 | 1.00 | 8.22 | 1.00 | 1.00 | 0.03% |
| 2d. MaxMV | 1.00 | 12.90 | 1.00 | 1.00 | 0.31 | 1.00 | 4.00 | 0.98 | 0.98 | 1.83% |
| 2e. Mask+Vac+MaxMV | 1.00 | 12.90 | 0.41 | 0.64 | 0.31 | 1.00 | 1.04 | 0.65 | 0.65 | 35.37% |
| 2f. Mask+Vac+MedMV | 1.00 | 12.90 | 0.41 | 0.64 | 0.51 | 1.00 | 1.71 | 0.82 | 0.82 | 18.15% |
| 2g. Mask+Vac+NatV | 1.00 | 12.90 | 0.41 | 0.64 | 0.82 | 1.00 | 2.74 | 0.94 | 0.94 | 6.44% |
| 2h. Mask+Vac+MaxMV+New Variant | 1.00 | 12.90 | 0.41 | 0.64 | 0.31 | 10.00 | 10.39 | 1.00 | 1.00 | 0.00% |
| 2i. Outdoor | 1.00 | 12.90 | 1.00 | 1.00 | 0.03 | 1.00 | 0.44 | 0.36 | 0.36 | 64.16% |
| 2j. Mask+Vac+Outdoor | 1.00 | 12.90 | 0.41 | 0.64 | 0.03 | 1.00 | 0.12 | 0.11 | 0.11 | 89.11% |
| 2k.Mask+Vac+Outdoor+NewVariant | 1.00 | 12.90 | 0.41 | 0.64 | 0.03 | 10.00 | 1.15 | 0.68 | 0.68 | 31.56% |

| **3. Scenario 3: Exposed Persons Speaking and Infectious Persons No Speaking** |  |  |  |  |  |  |  |  |  |  |
| --- | --- | --- | --- | --- | --- | --- | --- | --- | --- | --- |
| **Public Health Interventions (PHIs)** | **AFE** | **AFI** | **MF** | **VAF** | **VEF** | **CVF** | **Viral Load (Quanta)** | **p** | **p-ratio** | **Reduction in p due to PHIs** |
| 3a. Baseline | 2.82 | 1.00 | 1.00 | 1.00 | 1.00 | 1.00 | 2.82 | 0.94 | 1.00 |  |
| 3b. Mask | 2.82 | 1.00 | 0.41 | 1.00 | 1.00 | 1.00 | 1.15 | 0.68 | 0.73 | 27.31% |
| 3c. Vac | 2.82 | 1.00 | 1.00 | 0.64 | 1.00 | 1.00 | 1.80 | 0.83 | 0.89 | 11.30% |
| 3d. MaxMV | 2.82 | 1.00 | 1.00 | 1.00 | 0.31 | 1.00 | 0.87 | 0.58 | 0.62 | 38.03% |
| 3e. Mask+Vac+MaxMV | 2.82 | 1.00 | 0.41 | 0.64 | 0.31 | 1.00 | 0.23 | 0.20 | 0.22 | 78.39% |
| 3f. Mask+Vac+MedMV | 2.82 | 1.00 | 0.41 | 0.64 | 0.51 | 1.00 | 0.37 | 0.31 | 0.33 | 66.89% |
| 3g. Mask+Vac+NatV | 2.82 | 1.00 | 0.41 | 0.64 | 0.82 | 1.00 | 0.60 | 0.45 | 0.48 | 52.05% |
| 3h.Mask+Vac+MaxMV+NewVariant | 2.82 | 1.00 | 0.41 | 0.64 | 0.31 | 10.00 | 2.27 | 0.90 | 0.95 | 4.63% |
| 3i. Outdoor | 2.82 | 1.00 | 1.00 | 1.00 | 0.03 | 1.00 | 0.10 | 0.09 | 0.10 | 90.17% |
| 3j. Mask+Vac+Outdoor | 2.82 | 1.00 | 0.41 | 0.64 | 0.03 | 1.00 | 0.03 | 0.02 | 0.03 | 97.35% |
| 3k.Mask+Vac+Outdoor+NewVariant | 2.82 | 1.00 | 0.41 | 0.64 | 0.03 | 10.00 | 0.25 | 0.22 | 0.24 | 76.30% |
| **4. Scenario 4: Exposed Persons and Infectious Persons – Both Speaking** |  |  |  |  |  |  |  |  |  |  |
| **Public Health Interventions (PHIs)** | **AFE** | **AFI** | **MF** | **VAF** | **VEF** | **CVF** | **Viral Load (Quanta)** | **p** | **p-ratio** | **Reduction in p due to PHIs** |
| 4a. Baseline | 2.82 | 12.90 | 1.00 | 1.00 | 1.00 | 1.00 | 36.38 | 1.00 | 1.00 |  |
| 4b. Mask | 2.82 | 12.90 | 0.41 | 1.00 | 1.00 | 1.00 | 14.84 | 1.00 | 1.00 | 0.00% |
| 4c. Vac | 2.82 | 12.90 | 1.00 | 0.64 | 1.00 | 1.00 | 23.17 | 1.00 | 1.00 | 0.00% |
| 4d. MaxMV | 2.82 | 12.90 | 1.00 | 1.00 | 0.31 | 1.00 | 11.28 | 1.00 | 1.00 | 0.00% |
| 4e. Mask+Vac+MaxMV | 2.82 | 12.90 | 0.41 | 0.64 | 0.31 | 1.00 | 2.93 | 0.95 | 0.95 | 5.33% |
| 4f. Mask+Vac+MedMV | 2.82 | 12.90 | 0.41 | 0.64 | 0.51 | 1.00 | 4.81 | 0.99 | 0.99 | 0.81% |
| 4g. Mask+Vac+NatV | 2.82 | 12.90 | 0.41 | 0.64 | 0.82 | 1.00 | 7.73 | 1.00 | 1.00 | 0.04% |
| 4h. Mask+Vac+MaxMV+NewVariant | 2.82 | 12.90 | 0.41 | 0.64 | 0.31 | 10.00 | 29.31 | 1.00 | 1.00 | 0.00% |
| 4i. Outdoor | 2.82 | 12.90 | 1.00 | 1.00 | 0.03 | 1.00 | 1.25 | 0.71 | 0.71 | 28.61% |
| 4j. Mask+Vac+Outdoor | 2.82 | 12.90 | 0.41 | 0.64 | 0.03 | 1.00 | 0.33 | 0.28 | 0.28 | 72.24% |
| 4k. Mask+Vac+Outdoor+NewVariant | 2.82 | 12.90 | 0.41 | 0.64 | 0.03 | 10.00 | 3.25 | 0.96 | 0.96 | 3.87% |

**Table 4b.** E(I) for different activities and public health interventions, for fixed parameter values of group size N, duration T and ratio of infectious persons to total persons M/N

| **1. Scenarios for Exposed-Breathing and Infectious-Breathing Only** |  |  |  |  |  |  |  |  |  |  |
| --- | --- | --- | --- | --- | --- | --- | --- | --- | --- | --- |
| Public Health Interventions (PHIs) | AFE | AFI | MF | VAF | VEF | CVF | Viral Load (Quanta) | p | E(I) w. N=50 (500), M/N=0.3, T=1 | E(I) w. N=50 (500), M/N=0.5, T=1 |
| 1a. Baseline | 1.000 | 1.000 | 1.000 | 1.000 | 1.000 | 1.000 | 1.000 | 0.632 | 18.94 (186.38) | 18.17 (179.60) |
| 1e. Mask+Vac+MaxMV | 1.000 | 1.000 | 0.408 | 0.637 | 0.310 | 1.000 | 0.081 | 0.077 | 0.30 (2.93) | 0.35 (3.48) |
| **2. Scenarios for Exposed-Breathing and Infectious-Speaking Only** |  |  |  |  |  |  |  |  |  |  |
| 2a. Baseline | 1.000 | 12.90 | 1.000 | 1.000 | 1.000 | 1.000 | 12.90 | 1.000 | 24.8 (244.96) | 21.8 (216.37) |
| 2e. Mask+Vac+MaxMV | 1.000 | 12.90 | 0.408 | 0.637 | 0.310 | 1.000 | 1.039 | 0.646 | 19.21 (189.11) | 18.37 (181.55) |
| **3. Scenarios for Exposed-Speaking and Infectious-Breathing Only** |  |  |  |  |  |  |  |  |  |  |
| 3a. Baseline | 2.820 | 1.000 | 1.000 | 1.000 | 1.000 | 1.000 | 2.820 | 0.940 | 24.05 (237.09) | 21.40 (212.06) |
| 3e. Mask+Vac+MaxMV | 2.820 | 1.000 | 0.408 | 0.637 | 0.310 | 1.000 | 0.227 | 0.203 | 7.72 (75.82) | 8.49 (83.57) |
| **4. Scenarios for Exposed-Speaking and Infectious-Speaking Only** |  |  |  |  |  |  |  |  |  |  |
| 4a. Baseline | 2.820 | 12.90 | 1.000 | 1.000 | 1.000 | 1.000 | 36.39 | 1.000 | 24.8 (244.96) | 21.8 (216.37) |
| 4e. Mask+Vac+MaxMV | 2.820 | 12.90 | 0.408 | 0.637 | 0.310 | 1.000 | 2.931 | 0.947 | 24.15 (238.04) | 21.44 (212.59) |

Our results show that for non-speaking activities, the probability of infection p can be substantially reduced from the baseline scenario (Scenario 1a) by 87.75% when three PHIs – masking, three-jab vaccination, and maximum ventilation – are implemented (Scenario 1e). The same measures lead to similar reductions in p of 78.39% from the baseline scenario (Scenarios 3a, 3e) in situations where exposed persons speak but infectious persons do not speak.

However, the effectiveness of these PHIs is severely compromised when infectious persons speak. When exposed persons do not speak but infectious persons do, the reduction in p from the baseline scenario (Scenario 2a) drops to 35.37% (Scenario 2e), while when both exposed persons and infectious persons speak, the reduction from the baseline scenario (Scenario 4a) drops to 5.33% (Scenario 4e), such that neither individual nor a combination of public health interventions lead to any meaningful reduction in the probability of infection.

We also consider the effect of individual PHIs on the probability of infection. Their ordinal ranking is consistent across activities, with maximum mechanical ventilation (MaxMV) having the greatest effect on reducing p, followed by masking (Mask), and vaccination (VAC). As above, these have a meaningful effect only when infectious persons do not speak. When infectious persons speak, then even MaxMV reduces p by only 1.83% (Scenario 2d) from the baseline scenario (Scenario 2a) and when both infectious persons and exposed persons speak, then no PHI is effective (Scenarios 4a, 4d).

If variant infectivity (CVF) is increased ten-fold from Omicron, then even in the most favorable situation of infectious and exposed persons doing nothing but breathing, the strongest PHI combination (Mask plus Vac plus MaxMV) reduces p by just 12.48% (Scenario 1h), as compared to the Baseline (Scenario 1a) with no PHIs. In all other situations, PHIs cease to have any meaningful impact in reducing infections (Scenarios 2h, 3h, and 4h).

## 3. Discussion and Conclusions

### 3.1 Time spent indoors

For a given number of infectious persons M in a room with a total of N persons, the expected number of infected persons E(I) always increases with time T spent in the room. All other factors being equal, shorter gatherings are preferred.

The incremental increase in E(I) for an additional period varies with M/N. From Figures 1b-1d (reflecting p = 1), for T = 1, E(I) initially increases with M/N until E(I) peaks at M/N = 30% and then decreases for higher M/N. Dividing the curve into 3 sections – a left-hand section where E(I) is increasing, peak E(I), and a right-hand section where E(I) is decreasing – it can be seen that as T increases, the left-hand section shifts up and flattens, peak E(I) increases and shifts leftwards (i.e. occurs at lower values of M/N), and the right-hand section more rapidly approaches linear decline. As meetings become longer, there is less sensitivity as to whether a single infectious person or multiple infectious persons are initially present because the increase in exposure time tends towards saturation, such that everyone will be infected either way. This shifts policy emphasis; it is always desirable to have fewer infectious persons in a room but, whereas in shorter meetings there is benefit in reducing M/N, in longer meetings most benefit occurs from ensuring M/N = 0. As a consequence, longer meetings would benefit from more stringent control measures, such as prior PCR testing of individuals rather than rapid antibody tests (RAT), to ensure that M = 0, as well as smaller groups to limit the probability of a single infectious person entering the room – a risk that increases with group size.

Finally, the impact of N on E(I) is highly constrained which can be seen visually from the similarity across Figures 1b, 1c and 1d for N = 50, 100 and 500 respectively.

### 3.2 Group partitioning

The effect of group partitioning or restricting group sizes is very modest. Referring to Table 3a, we illustrate this for N = 100 and p = 0.5. For M/N = 30%, E(I) falls from 31.87 for 1 partition (PAR = 1) to 30.75 when the group is divided into 5 partitions (PAR = 5), i.e., an absolute decrease of 1.12 or relative decrease of 3.51%. Intuitively, the effect is limited because partitioning does not directly stop infection. Instead, the effect is achieved via reducing the probability of finding a new person to infect for a given total group size: more partitions mean fewer people within each partition and hence a reduced maximum number of people that a given infectious person can infect, so saturation within a partitioned group occurs more quickly.

For cases where the infected proportion of the group is low, reaching saturation is relatively unlikely, so partitioning has little effect. Higher p causes saturation to be more likely, increasing the effect of partitioning. Thus, for N = 100 and M/N = 10%, the relative decrease in E(I) for PAR = 1 to PAR = 5 is 1.12% for p = 0.1 but increases to 5.53% for p = 1.

For cases where the infected proportion of the group is already high, reaching saturation is relatively likely, so partitioning again has little effect. Since lower p causes saturation to be less likely, this increases the effect of partitioning. Thus, for N = 100 and M/N = 50%, the relative decrease in E(I) for PAR = 1 to PAR = 5 is 4.15% for p = 0.1 but decreases to 3.27% for p = 1.

Notwithstanding the limited effectiveness of restricting group size, the use of such measures should consider the phase of infection cycle and probability of infection. For example, for a highly infectious variant (p → 1), restricting group size is relatively more effective during the early phases of an outbreak, whereas for a less infectious variant (p → 0), restricting group size is relatively more effective during later phases.

In the real-world, restrictions on group size have been amongst the most widely implemented PHIs to control the pandemic. Several studies have suggested that such measures have been effective in reducing transmission, with restrictions on small groups being more effective than on large groups. For instance, Sharma *et al*. (2021) found that restricting gatherings to 2 people was considerably more effective than restricting gatherings to 10 people, which in turn was slightly more effective than restricting gatherings to 30 people, with stricter mask wearing policies being more effective than restricting gatherings to 10 people.^41^ Haug *et al*. (2020) found that the most effective PHI was cancelling small gatherings (including gatherings of up to 50 people, closures of shops and restaurants, and mandatory home working).^42^ There are several reasons for the apparent discrepancy between these findings and our model. Most importantly, the studies do not distinguish between frequency and per event impact: even a very modest effect repeated many times can have a considerable aggregate impact. Although M/N, the proportion of individuals infected early in an outbreak is generally small (<0.01), meetings seldom occur with social distancing in place, so there can be a large impact as infection spreads through sequential meetings at low N. Additionally, the restrictions themselves (especially restricting gatherings to 2 people) are likely to affect people’s behavior and reduce the number of gatherings (which is outside the scope of this model’s one-shot structure).

Real-world studies also suggest that the distribution of transmission follows an exponential distribution rather than a uniform distribution. For example, Laxminarayan *et al*. (2020) estimated that 5% of infected individuals accounted for 80% of cases in the Indian states of Tamil Nadu and Andhra Pradesh and Adam *et al*. (2020) found that 19% of cases caused 80% of local transmissions in Hong Kong.^43,44^ “Super-spreaders” likely reflect a combination of biological, behavioral and environmental factors. Interpreted in the framework of this model, super-spreaders may exhibit much greater viral shedding (it has been suggested that individuals infected by Omicron may exhale levels of virus 1000 times higher than the levels of Alpha or Delta strains^45^) and be part of larger and more numerous partitions than typical individuals. Hence, earlier interventions that restrict group size N and movement (for example, geographically isolating areas or quarantining), may act to mitigate such cases and significantly reduce transmission, though relative effectiveness may decline as infection rates increase.

### 3.3 Speaking interactions

Table 4a shows that speaking has a large effect on the probability of infection p, especially if infectious persons speak. In the baseline scenario in which neither infectious persons nor exposed persons speak (Scenario 1a), viral load quantum = 1 (by definition) and p = 0.63. By comparison, viral load = 2.82 and p = 0.94 when only exposed persons speak (Scenario 3a) and viral load = 12.90 and p = 1.00 when only infectious persons speak (Scenario 2a). Since the effect of exposed persons speaking and the effect of infectious persons speaking interact multiplicatively, when both exposed and infectious persons speak (Scenario 4a), viral load = 36.38 and p = 1.00.

Under maximum PHI (masking, vaccination, and maximum ventilation), when both infectious persons and exposed persons speak (Scenario 4e), viral load is reduced to 2.93 and p is reduced to 0.95. The reduction in viral load is much larger than the reduction in infection probability, reflecting the viral load being far above the minimum needed to cause infection. This model treats infection as a binary variable but in practice there may be benefits to lowering the viral load, though the relationship between viral load and infection probability-severity is not yet definitively understood.^12^

For Omicron, infectious persons speaking essentially negates maximum PHI (viral loads and infection probability under Scenario 3e are similar to those of no speaking and no PHI under Scenario 1a) and both infectious and exposed persons speaking overwhelms maximum PHI (viral loads are approximately 3 times higher and probability of infection is approximately 50% higher under Scenario 4e than no speaking and no PHI under Scenario 1a). This motivates widespread frequent Rapid Antigen Testing (RAT). Even with sensitivity rates of 70%, RAT and self-isolation (with considerably less than 100% compliance) is likely to mitigate infection more than maximum levels of PHI in conversational settings (p declines from 1.00 with no PHI under Scenario 4a to 0.95 with maximum PHI under Scenario 4e). If public health authorities seek to control the spread of SARS-CoV-2, this may be the only alternative to remote living. The scope and appeal of restricting in-person conversation seems limited; conversation is intrinsic to human society and the nature and purpose of most in-person gatherings. Modern society is ill-adapted for monastic silence. For the relatively few settings in which communication is naturally one-way – for example, meet-and-greet receptions and lecturers/teachers in lecture halls/classrooms – it seems prudent for the speaking individuals to take additional precautions given the strong asymmetrical effect of an infectious person speaking, but as a policy, it seems highly questionable whether PHIs to permit gatherings provided verbal communication is one-way would be broadly practical or even desirable (in many cases, this would defeat the point of gathering).

### 3.4 Public health interventions and consideration of outdoor gatherings and potential future high-infectivity variants

As PHIs – masking, vaccination, and ventilation – combine multiplicatively, the greatest reduction in probability of infection p comes when all three measures are introduced. The most effective individual PHI is maximum ventilation (MaxMV), followed by wearing masks and then by vaccination. When there is no speaking (Scenario 1), baseline infection probability is 0.632 (Scenario 1a), falling to 0.077 under maximum PHI (scenario 1e), 0.267 under MaxMV (Scenario 1d), 0.335 under masking (Scenario 1b), and 0.471 under vaccination (Scenario 1c). As discussed previously, all interventions are greatly affected by speaking but this ordinal ranking is preserved (Scenarios 2-4). In the Supplementary Materials, we consider the threshold value of p such that exponential growth is avoided early in an infection cycle; for the assumptions here (notably T = 1), threshold p is approximately 0.3.

Improving ventilation for indoor environments warrants greater public health attention. This reduces infection probability by more than face masks or vaccinations and its effect is incremental to other PHIs. A similar alternative is the use of air filtration systems, such as portable High Efficiency Particulate Air (HEPA) cleaners,^12^ though efficacy will depend on ventilation in the sense of air in the room being well mixed. Addressing ventilation is also likely less contentious, since this avoids many issues related to not wearing face masks or being vaccinated, and becomes even more important in the absence of masking or vaccination.

Switching from indoors venues to outdoors (Outdoor) has an effect similar to the impact of conversing. Baseline infection probability with no speaking and no PHI is 0.632 (Scenario 1a); with speaking and no PHI, infection probability increases to 1.00 (Scenario 4a); but with speaking and meeting outdoors, infection probability falls to 0.714 (Scenario 4i). With face masks and vaccination, meeting outdoors further reduces infection probability to 0.278 (Scenario 4j). Clearly, this is subject to weather and climate but when and where appropriate, for example in sub-tropical/tropical regions or in warmer months in temperate regions, moving outdoors is a quick, effective and low-cost measure.

Face masks are relatively affordable and now widely available but have met with resistance from a significant proportion of Western society and mandatory masking may be sufficiently publicly unpopular as to prevent the re-introduction of such measures.

Vaccines play a crucial role in mitigating the severity of illness but are less effective in preventing infection relative to other measures. Additionally, in the developing world, the cost of vaccines and limited supporting infrastructure mean vaccination may not be an option, whilst in Western countries, there are significant segments of society unwilling to be vaccinated.

Lastly, we observe that a hypothetical new variant with infectivity an order of magnitude greater than Omicron would pose serious challenges to PHIs. In a no speaking setting, Omicron has an infection probability of 0.63 with no PHI (Scenario 1a), whilst this hypothetical new variant would have an infection probability of 0.55 under maximum PHI (masking, vaccination, and maximum ventilation; Scenario 1h), assuming the practical challenges discussed above can be overcome. But if both infectious persons and exposed persons speak, then even moving outdoors will be insufficient (p = 0.96 under Scenario 4k).

### 3.5 Limitations

Practical application of the model would benefit from refining parameterization and the introduction of additional parameters, for example to distinguish between different types of masks or vaccines. The model structure can be extended to more accurately capture real-world dynamics. One important dimension is heterogeneity, for example to reflect different levels of contagiousness and susceptibility as well as non-uniform population distancing and contact times. Additionally, the model could be extended from a “one-shot” structure, which ends when people leave a room, to a “multi-shot” structure, which considers people leaving a room and entering new rooms.

### 3.6 Conclusions

We have proposed a multinomial probabilistic model to capture infection dynamics on a bottom-up individual basis. This helps to make the model relatively straightforward to understand and easy to adjust for variations in parameters or additional parameters. As discussed in the Limitations section, this framework can be further developed but we believe it can provide policy-makers with quick and intuitive guidance to assist in scenario planning.

This model underscores the immense practical challenges of eliminating SARS-CoV-2, as high SARS-CoV-2 infectivity greatly raises the cost of allowing even a single infectious person to come into contact with uninfected individuals. In this regard, whilst frequent mass testing and self-isolation can help to control outbreaks, as numbers increase, the probability of avoiding such instances may become untenably small.

Improving ventilation – whether by making physical alterations, moving meetings outdoors or using air filtration systems – should be given much greater attention than at present. Compared to other public health interventions, ventilation can have a larger impact on reducing infection, is incrementally beneficial (not mutually exclusive), and is likely to be less challenged in implementation. This is particularly relevant as societies seek to resume normal human interaction while controlling the spread of SARS-CoV-2, as our model also suggests that in-person conversation has an order of magnitude impact on infection probability (in scenarios where this increase is not constrained by the new infection probability approaching 1) and on viral load.

Finally, our model suggests partitioning groups has a limited impact on reducing transmission within a gathering. To the extent that studies suggest restrictions on group size have had an impact, this suggests a large hidden cost since the channel is not via reduced spread per event (as commonly perceived) but rather by affecting the number of gatherings.

## 4. Methods

### 4.1 Basic model: M infectious persons in a room with N total persons, with each infectious person making L contacts at random in a time period T, and infecting each exposed (healthy) person with probability p for each contact

Suppose we have a room of N persons, M infectious persons, and N-M exposed (healthy) persons, labelled 1, 2, …, N-M. The M infectious persons circulate among the N persons in a room, conducted over a duration T, and each infectious person will make L contacts with the others in the room at random, with the possibility of meeting another infectious person or with an exposed person more than once. Assuming infectious individuals will infect any exposed person with probability p each time a contact is made, what is E(I), the expected number of I, the number of exposed persons infected?

According to the US CDC^25^, individuals who are within six feet of an infectious person, via a face-to-face contact, for a total of 15 minutes or more, will be at risk of infection. Hence, we selected 15 minutes as a time of interest. Hence, in a 1-hour meeting event every infectious person entering a room will have an opportunity of contacting four persons. Therefore, we assume if one infectious person moves around the room and talks to different people freely, at random, every 15 minutes, he will make four personal contacts. The infectious person may talk to the same person in the room repeatedly. We assume that the infectious person will infect each person with a probability p for every single personal contact, lasting for about 15 minutes.

#### 4.1.1 Expected number of exposed subjects infected

Now we compute the probability of no one being infected after one contact. This single contact is equally likely to be caused by any of the M infectious persons, i.e., it is caused by infectious person 1 with the probability 1/M. The infectious person 1 will cause zero infection if he/she meets another infectious person, with the probability (M-1)/(N-1), or if he/she meets an exposed (healthy) person, with probability (N-1-(M-1))/(N-1), and does not infect the exposed person, with the probability (1-p). So the infectious person 1 not causing any infection is given by (M-1)/(N-1) + (N-M)/(N-1) × (1-p). Due to the independence between different infectious persons, the infectious person 2, 3, …, M will not cause any new infection with the same probability. So in one contact the probability that no new infection is caused is as follows:

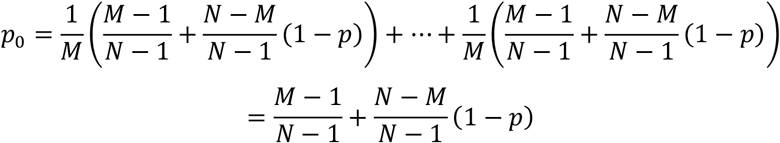

The probability of each of the N-M exposed (healthy) persons being infected should be the same as follows:

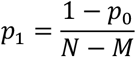

Then we can use a multinomial distribution to determine how many of those exposed to SARS-CoV-2 are getting productively infected. In each of the ML contacts, we have N-M+1 outcomes: Exposed person 1 gets infected with probability *p*_1_, exposed person 2 gets infected with probability *p*_1_,…, exposed person (N-M) gets infected with probability *p*_1_, whilst no one gets infected with probability *p*_0_. Note that here we consider only the infections obtained via this contact, without considering whether the exposed person has gotten infected before or not.

Next, we have a multinomial (*ML, p*_0_, *p*_1_, …, *p*_1_) distribution. Let *X*_*j*_ be the number of times out of the ML contacts that no one gets infected, *X*_0_ be the number of times the exposed person j is infected, for j = 1,…, N-M. Then we have

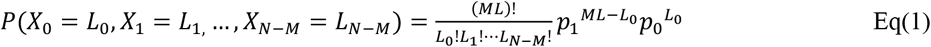

Now the number of exposed persons infected is ∑_*j*≠0_ *Ind*(*L*_*j*_ > 0) where *Ind*() is the indicator function. The expected number of people getting infected is:

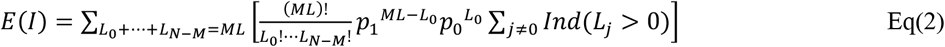

Now we simplify Eq(2) for computational purpose. We separate the situations according to the different values of *I*. Let *P*^*i*^ *=* ℙ(*I=i*).

First, consider the situation, when *L*_1_ = ∆ = *L*_*N*−*M*_ = 0. There is only one permutation, i.e., *L*_0_ = *ML, L*_1_ = ∆ = *L*_*N*−*M*_ = 0 that satisfies this condition. From Eq(1),

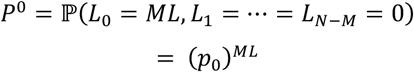

Second, consider the situation, when *L*_0_ = *ML* − *j, L*_1_ = *j, L*_2_ = ∆ = *L*_*N*−*M*_ = 0, From Eq(1), 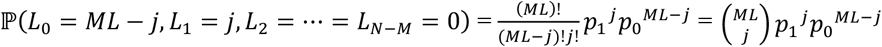

Hence, summing over the possible values of *j*,

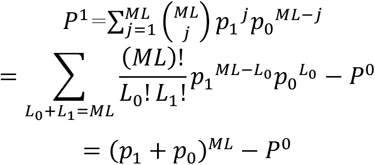

Then, given that ℙ(*L*_*k*_ ≠ 0 *and L*_j_ = 0 *for j* ≠ *k*) = *P*_1_ for all *k*s.

Now, consider the situation, when *L*_1_ ≠ 0, …, *L*_*k*_ ≠ 0, *L*_*k*+1_ = ∆ = *L*_*N*−*M*_ = 0,

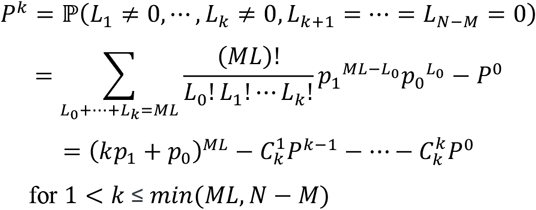

If *ML* < *N* − *M*, at most ML people will get infected, hence, *P*^*j*^ = 0 for *j* > *ML*.

The expected infection number is:

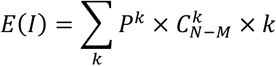

#### 4.1.2 Partition model: M infectious persons in one single group gathering of N persons, partitioned at random into R multiple rooms

When computing the expected number of exposed subjects infected for the partition model, we assume that each of the M infectious persons are equally likely to be in each of the R rooms and enumerate the different cases of how the M infectious persons are distributed in these R rooms. For example, suppose there are 5 rooms, the probability of each infectious person in each room is 1/5. If the number of infectious people is 10, then the probability that all the 10 infectious people are in one room is 5 × (1/5)^10^ = 1/5^9^.

To compute the expected number of exposed subjects infected, we compute the probabilities of all the enumerated cases, and the expected number of exposed subjects infected in each case. Then we can use the law of total expectation to get the result.

#### 4.1.3 Probability of infection p

Our goal is to find the probability p that an exposed person will get infected when he/she is in contact with an infectious person. We want to factor in different scenarios/public health measures, such as (1) the activities conducted by the exposed person, and the infectious person, (2)masking, (3) vaccination, (4) ventilation, and (5) variant infectivity.

We start with Eq(3), developed in Buonanno *et al*.^15^, using a modified Wells-Riley equation:

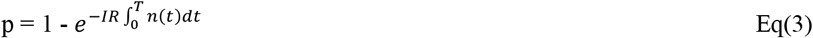

where IR = inhalation rate of the exposed person (m^3^/h)

n(t) = number of quanta/m^3^ at time t

T = time of exposure (h)

For relatively short exposure times, for instance, a few hours, we assume that n(t) is a constant n, and Eq(3) can be reduced to:

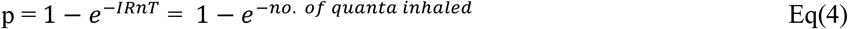

From Eq(4), we can see that, by setting the number of quanta inhaled as one, one virus quantum is the dose of airborne droplet nuclei required to cause infection in 1 – *e*^−1^ = 63% of exposed persons.

We can use viral quanta as the variable to study different scenarios of interest by properly modifying *IR* and *n. IR* will increase with the activity level of the exposed, while *n* (viral quanta/m^3^ in the air) will increase with the activity level of the infectious person and decrease when the infectious person is vaccinated. *n* will also decrease with ventilation and air filtering. Masking by either the exposed or the infectious person will also decrease the effective *n* as some of the virus will be blocked by masking.

Hence Equ. (4) can be modified as follow:

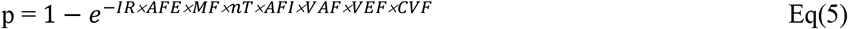

where *IR* has been modified by *AFE*, the activity factor of the exposed, while *n*, the number of viral quanta emitted by the infectious person is modified by *AFI*, the activity factor of the infectious person, *VAF*, the vaccination factor of the infectious person, *VEF*, the ventilation and filtering factor, and *CVF*, the SARS-CoV-2 variant factor. *MF* is the masking factor.

Next we study how to determine each of these modifying factors:

##### AFE

Buonanno *et al*. (2020)^15^ found the inhalation rate of five different activities of the exposed in an indoor setting, namely, resting, standing, light exercise, moderate exercise, and heavy exercise, averaged between males and females, to be 0.49, 0.54, 1.38, 2.35, and 3.30 m^3^/h, respectively. Thus, if we take resting as the baseline (with an *AFE* of 1), then the *AFE* for the five activities will be 1, 1.10, 2.82, 4.80, and 6.73, respectively. In Table 4, we assume the activity of speaking is similar to light exercise.

##### AFI

Coleman *et al*. (2021)^26^ compared the different viral loads emitted by the infectious person when they are breathing, talking, and singing, and found viral loads of 1960, 25312, and 32831 virus gene copies, in a period of 30 minutes. Coleman *et al*. (2022) actually reported the viral loads for the latter two activities for a period of 15 minutes each. To match the value reported for the activity of breathing for 30 minutes, we doubled the reported values to get the 30-minute values. If we take breathing as the baseline, with AFI =1, then the AFI for speaking and singing will be 12.9 and 16.8.

##### MF

Talic *et al*. (2021)^46^ surveyed 72 studies that assessed the effectiveness of public health measures in reducing the incidence of SARS-CoV-2 transmission, and SARS-CoV-2 mortality, and based on data of 35 of such studies, which evaluated the effect of individual health measures, rather than a package of measures, estimated that the reduction of the incidence of SARS-CoV-2 with masking is 0.47. Taking no masking as the baseline, MF for masking is 0.408, which is obtained as the ratio of the estimated viral quanta with masking and without masking.

##### VAF

Accorsi *et al*. (2022)^47^ estimated the C_t_ values for the Delta and Omicron variants of SARS-CoV-2, and found that for the Delta variant, the C_t_ values for unvaccinated and vaccinated (three doses) were 18.28 and 19.07, respectively. The corresponding numbers were 18.71 and 19.35 for the Omicron variant. Singanayagam et al.^11^ quoted a conversion equation between C_t_ and viral load, published by Public Health England, as follow:

Viral load (copies /ml) = 133e^(37.933 - Ct)/1.418^

If the unvaccinated is the baseline, with a *VAF* of 1, then we can easily calculate the *VAF* for vaccinated infectious person as 0.573 for the Delta variant, and 0.637 for the Omicron variant.

##### VEF

After a detailed study of the literature covering ventilation rates and SARS-CoV-2 infections in the US and EU/UK, we have decided to focus our ventilation setting on the classroom setting within the US (given the variation in ventilation rates over different environmental settings and building codes in different countries). ACH_T_ is used as the ventilation metric, defined as the rate at which room air is replaced by recirculated and outdoor air as determined by measuring the decay of a tracer species (such as CO_2_). From McNeill et al. (2022)^48^, we obtained the values for maximum mechanical ventilation rate (MaxMV) at ACH_T_ 8.7, minimum mechanical ventilation rate (MinMV) 2.7, median ventilation rate (MedMV) 5.3, and natural ventilation rate (NatV) 3.3 for the school classroom setting based in the US. Using the MinMV of 2.7 as the baseline, we can calculate the *VEF* for MaxMV, MedMV, and NatV as 0.310, 0.509, and 0.818, respectively. For outdoor ventilation (Outdoor), based on a thorough review of reported Covid-19 cases by Bulfone *et al*. (2021)^38^, the odds of transmission indoors are 18.7 times higher than outdoors. Using Eq(5), the ratio of Viral Load or quantum, Q, between Outdoor and Indoor, is 0.0344, which is derived from the following equations:

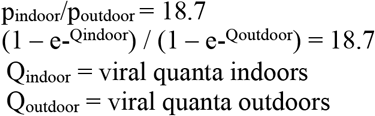

Using baseline of Q_indoor_ = 1 virus quantum, we have:

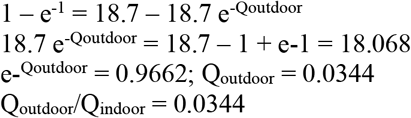

##### CVF

It is well-known that different SARS-CoV-2 variants will have different levels of infectivity. A recent study by the University of Hong Kong^17^ found that that Omicron infects and multiplies 70 times faster than the Delta variant and the original coronavirus in the human bronchus, which may explain why Omicron may transmit faster between humans than previous variants. Thus, if the baseline of Omicron is assigned a CVF of 1, then the CVF of Delta will be 1/70. In the future, we may have variants that are even more infectious than Omicron, in which case the CVF of such a variant will be bigger than 1.

Based on these factors, we can compute p-ratio, the ratio of p compared to the baseline, for four sets of scenarios, as summarized in Table 4 above.

## Data Availability

All data used in this study are included in the article.

## Acknowledgements

This research is supported in part by the Theme-based Research Scheme of the Research Grants Council of Hong Kong, under Grant No. T41-709/17-N.

## Data Availability

All data used in this study are included in the article.

## Code Availability

No custom code was used in the data analysis.

## Author contributions statement

VL and JL were jointly responsible for full conceptualization and methodological design of a generalized multinomial probabilistic model for SARS-CoV-2 infection prediction, to cater for public health policy intervention evaluation; wrote the first draft; and acquired research funding. YS contributed partially to literature review, refined the model and performed computations. YH contributed partially to the draft in relation to the sensitivity of the results to infection probability, and suggested computational simplifications. KC edited the draft and contributed partially to the discussions and conclusions. SW assisted in reference formatting, contributed partially to literature review, and proposed the Wells-Riley equation for infection risk assessment. JC observed that ventilation is one of the most significant factors driving COVID infection, and this inspiration has guided some of the key arguments in this paper. JD inspired us to frame the risk of infection based on viral loads, and contributed partially to the discussion and the abstract. QZ performed computations based on the model developed by VL.

## Additional information

The authors declare that they have no known competing financial interests or personal relationships that could have appeared to influence the work reported in this paper.

