## Supplementary Materials for "A Generalized Multinomial Probabilistic Model for SARS-CoV-2 Infection Prediction and Public Health Intervention Assessment in an Indoor Environment"

Victor OK Li[1,#,*], Jacqueline CK Lam[1,#,*], Yuxuan Sun[1], Yang Han[1], Kelvin Chan[1], Shan-shan Wang[1], Jon Crowcroft[2], Jocelyn Downey[1], Qi Zhang[1]
[1]Department of Electrical and Electronic Engineering
The University of Hong Kong, Hong Kong
[2]Department of Computer Science and Technology
The University of Cambridge, Cambridge, UK

[#]Equal Contributions
*Correspondence Authors

### The threshold of infection probability $p$ for epidemic control

While the infection probability $p$ can be calculated by factoring in different scenarios and public health interventions, for epidemic control and health policymaking, it is also critical to find a threshold of $p$ that gives plausibly slow growth in infected persons, such that the wave of infections can be easily controlled by additional interventions. The basic reproduction number, $R_0$, is widely used to measure disease spread and suggest control strategies.[1] For the current study, the event reproduction number, $R_{event}$, is utilized to determine the threshold of $p$, given that $R_{event}$ is more suitable for epidemic management in indoor spaces and across different settings of interventions.[2,3] $R_{event}$ is defined as the expected number of newly infected persons at an event due to the attendance of a single infectious person and can be calculated using the following procedure. First, based on the calculation procedure of Eq(2) and the linearity of expectation, for the basic model without group partitioning, $E(I)$ is calculated in closed form as follows:

$$E(I) = E\left(\sum_{j=1}^{N-M} Ind(L_j > 0)\right)$$
$$= \sum_{j=1}^{N-M} E\left(Ind(L_j > 0)\right) = \sum_{j=1}^{N-M} \mathbb{P}(L_j > 0)$$
$$= \sum_{j=1}^{N-M} \left(1 - \mathbb{P}(L_j = 0)\right) = \sum_{j=1}^{N-M} (1 - (1 - p_1)^{ML}) = (1 - (1 - p_1)^{ML})(N - M)$$

where $I$ represents the number of exposed (healthy) persons that are infected, $N$ is the total number of persons in the room, $M$ is the number of infectious persons, $L_j$ represents the number of times exposed person $j$ is infected, $Ind$ is an indicator function (the value is 1 if $L_j > 0$, otherwise 0), $L$ is the number of random contacts each infectious person makes with the others in the room (during a period of $T$), and $p_1$ is the probability of each of the $N - M$ exposed (healthy) persons being infected (see Section 4.1.1 in the main text).

Then, when $M = 1$ (i.e., a single infectious person), $R_{\text{event}}$ is calculated as follows:

$$R_{\text{event}} = \left(1 - \left(1 - \frac{p}{N-1}\right)^L\right)(N - 1)$$

where $p$ is the infection probability.

When group partitioning is implemented, the single infectious person is equally likely to be in each group partition. Therefore, based on the law of total expectation, $R_{\text{event}}$ is further calculated as follows:

$$R_{\text{event}} = \sum_{k=1}^{K} \left(E(I|\text{infectious individual in } k) \times \frac{1}{K}\right) = \left(1 - \left(1 - \frac{p}{N_{\text{PAR}} - 1}\right)^L\right)(N_{\text{PAR}} - 1)$$

where $k$ represents one group partition (room), $K$ is the total number of group partitions (rooms), and $N_{\text{PAR}}$ is the group partition size (i.e., $N/K$).

Next, given $L > 0$, $N_{\mathrm{PAR}} > 1$, and $\frac{p}{N_{\mathrm{PAR}}-1} < 1$, according to the two inequalities, namely, $1 + rx \leq (1+x)^r \leq e^{rx}$ (for $x > -1$ and $r > 0$), an upper bound and lower bound of $R_{\mathrm{event}}$ are calculated as follows:

$$\left(1 - e^{\frac{-pL}{N_{\mathrm{PAR}}-1}}\right)(N_{\mathrm{PAR}} - 1) \leq R_{\mathrm{event}} \leq \left(1 - \left(1 - \frac{pL}{N_{\mathrm{PAR}} - 1}\right)\right)(N_{\mathrm{PAR}} - 1) = pL$$

Finally, based on the inequality, namely, $\ln(x) \leq x - 1$, an upper bound and lower bound of the infection probability $p$ are defined by the event reproduction number as follows:

$$\frac{R_{\mathrm{event}}}{L} \leq p \leq \frac{(N_{\mathrm{PAR}} - 1)}{L} \ln\left(\frac{N_{\mathrm{PAR}} - 1}{N_{\mathrm{PAR}} - 1 - R_{\mathrm{event}}}\right) \leq \frac{N_{\mathrm{PAR}} - 1}{N_{\mathrm{PAR}} - 1 - R_{\mathrm{event}}} \frac{R_{\mathrm{event}}}{L}$$

The upper bound derived from the proposed model is consistent with previous epidemic modeling. Based on a standard SIR epidemic model, the basic reproduction number $R_0$ can be calculated by the product of transmission rate, contact rate, and infection duration (while assuming constant rates over time).[4] As a result, $R_0 = pL$. Moreover, when $N_{\mathrm{PAR}} \to \infty$, $p \sim \frac{R_{\mathrm{event}}}{L}$ and $R_{\mathrm{event}} \sim pL$, suggesting that when the population becomes larger, the $R_{\mathrm{event}}$ derived from the proposed model is asymptotically-equivalent to the $R_0$ derived from the SIR epidemic model. This result also suggests that compared to the proposed model, the SIR model may overestimate the disease reproduction number in small groups in indoor spaces.

To prevent an epidemic characterized by the exponential growth in infected persons at the initial stage of an infection cycle, the basic reproduction number $R_0$ should be lower than one.[1] In general, the event reproduction number $R_{\mathrm{event}}$ is no larger than the actual basic reproduction number $R_0$ due to the exposure to one-shot or multi-shot event, i.e., $R_{\mathrm{event}} \leq R_0 < 1$.[2] Therefore, the threshold of infection probability $p$ (the maximum acceptable $p$ to prevent an epidemic from exponential growth) is as follows:

$$p_{\mathrm{threshold}} = \left(1 + \frac{1}{N_{\mathrm{PAR}}-2}\right) \times \frac{1}{L} = \left(1 + \frac{1}{N_{\mathrm{PAR}}-2}\right) \times \frac{1}{4T}$$

Generally, the smaller the number of persons in a room, the higher the maximum acceptable *p*. The shorter the duration time in a room, the higher the maximum acceptable *p*. When PAR=1, T=1, for N=50, 100, and 500, the threshold of *p* is 0.255, 0.253, and 0.251, respectively. When PAR=5, T=1, for N=50, 100, and 500, the threshold of *p* is 0.281, 0.264, and 0.253, respectively. These threshold values can help evaluate when and where interventions are effective in controlling the growth of infected persons during the early stage of an infection cycle. Suppose we assume that the infection probabilities under different interventions and scenarios in Table 4 can reflect the actual situation in the real world. In that case, the combination of face masks, vaccination, and natural ventilation is sufficient to prevent an epidemic in Scenario 1 (where both exposed and infectious persons do not speak). However, when both exposed and infectious persons speak (Scenario 4), changing meeting venues from indoors to outdoors becomes necessary to stop the spread of infection. Nevertheless, if a hypothetical new variant with higher infectivity were introduced, increasing ventilation by moving outdoors would be insufficient in Scenario 4.
